# SARS-CoV-2 specific cellular and humoral immunity after bivalent BA.4/5 COVID-19 vaccination in previously infected and non-infected individuals

**DOI:** 10.1101/2023.05.03.23289472

**Authors:** Rebecca Urschel, Saskia Bronder, Verena Klemis, Stefanie Marx, Franziska Hielscher, Amina Abu-Omar, Candida Guckelmus, Sophie Schneitler, Christina Baum, Sören L. Becker, Barbara C. Gärtner, Urban Sester, Marek Widera, Tina Schmidt

**Author notes:** Correspondence: Martina Sester, PhD, Saarland University, Department of Transplant and Infection Immunology, Center for Infectious Diseases, Building 77, Kirrberger Straße, D-66421 Homburg, Germany.

## Abstract

Knowledge is limited as to how prior SARS-CoV-2 infection influences cellular and humoral immunity after booster-vaccination with bivalent BA.4/5-adapted mRNA-vaccines, and whether vaccine-induced immunity correlates with subsequent infection. In this observational study, individuals with prior infection (n=64) showed higher vaccine-induced anti-spike IgG antibodies and neutralizing titers, but the relative increase was significantly higher in non-infected individuals (n=63). In general, both groups showed higher neutralizing activity towards the parental strain than towards Omicron subvariants BA.1, BA.2 and BA.5. In contrast, CD4 or CD8 T-cell levels towards spike from the parental strain and the Omicron subvariants, and cytokine expression profiles were similar irrespective of prior infection. Breakthrough infections occurred more frequently among previously non-infected individuals, who had significantly lower vaccine-induced spike-specific neutralizing activity and CD4 T-cell levels. Thus, the magnitude of vaccine-induced neutralizing activity and specific CD4 T-cells after bivalent vaccination may serve as a correlate for protection in previously non-infected individuals.

## Introduction

SARS-CoV-2 variants of concern (VOCs) such as Omicron (B.1.1.529) have shown increased escape from neutralizing antibodies, which reduces the ability to prevent infection^1–3^. Together with waning immunity, this led to a substantial increase in the incidence of SARS-CoV-2 infections^4^. As a result, bivalent mRNA booster vaccines encoding the spike proteins of the ancestral WA1/2020 strain and of either the Omicron BA.1 or BA.5 sublineages have been developed^5, 6^. Most available immunogenicity studies have largely been restricted to neutralizing antibody activity^6–10^. Neutralizing antibody titers were shown to increase after vaccination with the bivalent vaccine to a slightly larger or similar extent as after monovalent vaccination, and the titers against the ancestral strain remained higher than against the Omicron strains^7–9^.

Despite the increase in infections in the Omicron era, the incidence of severe disease remained considerably low in otherwise healthy individuals^1, 2, 11^. This may be due lower virulence of the Omicron subvariants, to an increasing number of individuals with hybrid immunity and/or to the fact that SARS-CoV-2 specific T-cells, which have been shown to mediate protection from severe disease^12, 13^, are less affected by mutations in the VOCs spike protein. Up to now, knowledge on the induction of spike-specific CD4 and CD8 T-cells and on the impact of previous infection on immunogenicity after bivalent vaccination is limited, as most studies have reported aggregated data with small sample sizes^8, 9^. As this knowledge may have implications for future vaccine recommendations, this observational study was set up to characterize spike-specific IgG, neutralizing activity, and CD4 and CD8 T-cells against the ancestral spike and the Omicron subvariants before and after bivalent vaccination in individuals with and without prior infection. So far, most studies have focused on humoral immunity as a correlate of protection, whereas much less attention has been given to vaccine-induced cellular immunity^14^. Therefore, all study participants were followed up to associate individual vaccine-induced antibody and T-cell levels with the ability to protect from subsequent breakthrough infection.

## Results

### Study population

We recruited 127 immunocompetent individuals (49.5±13.5 years) who underwent COVID-19 vaccination with a bivalent COVID-19 vaccine (Comirnaty Original/Omicron BA.4/5, BioNTech/Pfizer), as per German regulations. Among them, 64 were grouped as previously infected either by self-reported history of SARS-CoV-2 infection (confirmed by PCR or rapid antigen test) and/or by a positive nucleocapsid protein (NCP) serology (“hybrid immunity group”). Sixty-three individuals were grouped as non-infected based on self-reporting and negative NCP-serology (table 1, “non-infected group”). Individuals either had 3 or 4 immunization events (mostly 3 vaccinations with and without 1 infection) before receiving the bivalent vaccine. An overview of the study design is shown in Extended data figure 1. Most individuals had received prior vaccinations with an mRNA vaccine, and a minor part had a history of heterologous vector/mRNA vaccinations. Individuals with prior infection were younger, and most infections had occurred in the Omicron BA.2 era. Blood samples were drawn before and 14 (IQR 3) days after vaccination to determine differential blood counts and vaccine-induced humoral and cellular immunogenicity. Demographic characteristics and differential blood counts are shown in table 1. General leukocyte and lymphocyte numbers did not differ between previously infected and non-infected individuals, except for monocytes which were significantly lower among individuals with prior infection (Table 1).

**Table 1:** Demographic and clinical characteristics of the study population.

|  | Infected<br>n=64* | Non-infected<br>n=63 | p-value |
| --- | --- | --- | --- |
| **Years of age [mean±SD]** | 46.5±13.7 | 52.6±12.7 | 0.011<sup>‡</sup> |
| **Sex, n (%)** |  |  | 0.090<sup>†</sup> |
| male | 26 (40.6) | 16 (25.8) |  |
| female | 38 (59.4) | 47 (74.6) |  |
| **Vaccine regimen** |  |  | 0.473<sup>†</sup> |
| mRNA | 40 (62.5) | 35 (55.6) |  |
| Vector/mRNA combination | 24 (37.5) | 28 (44.4) |  |
| **Number of immunization events (n)<sup>#</sup>** |  |  | <0.0001<sup>†</sup> |
| 3 | 5 (7.8) | 55 (88.7) |  |
| 4 | 59 (92.2)* | 8 (12.7) |  |
| **Analysis time [days after vaccination], median (IQR)** | 14 (2) | 15 (4) | 0.109<sup>‡</sup> |
| **Differential blood cell counts [cells/μl], median IQR** | n=63 | n=60 |  |
| Leukocytes | 6650 (2375) | 7100 (2900) | 0.134<sup>‡</sup> |
| Granulocytes | 4017 (1888) | 4351 (2350) | 0.166<sup>‡</sup> |
| Monocytes | 513 (192) | 573 (317) | 0.008<sup>‡</sup> |
| Lymphocytes | 2011 (766) | 2086 (831) | 0.977<sup>‡</sup> |
| Thrombocytes | 264000 (89000) | 293000 (82000) | 0.147<sup>‡</sup> |
| **Dominant SARS-CoV-2 strain at infection prior to vaccination<sup>§</sup>** |  |  |  |
| Parental | 5 (7.7) |  |  |
| Alpha | 1 (1.6) |  |  |
| Delta | 2 (3.1) |  |  |
| Omicron | 52 (80.0) |  |  |
| BA.1 | 7 |  |  |
| BA.2 | 41 |  |  |
| BA.4/5 | 4 |  |  |
| Unknown* | 4 (6.3) |  |  |

<sup>‡</sup>Unpaired t-test. <sup>†</sup>Fisher test; <sup>‡</sup>Mann-Whitney test; <sup>#</sup>immunization events (including infection(s) or vaccinations) prior to bivalent vaccination; <sup>§</sup>based on dominance of SARS-CoV-2 strain at the time of individual infection; \*includes 4 individuals without known history of infection, but positive NCP-ELISA.

### Adverse events after bivalent vaccination

Adverse events were analyzed in the first week after the bivalent vaccination based on self-reporting using a questionnaire. Approximately 80% of individuals reported local or systemic adverse events or both (figure 1A) with no differences between individuals with and without prior infection (p=0.872). Adverse events were overall mild, and pain at the injection site followed by fatigue were most frequently reported (figure 1B). Based on individual perception of adverse events compared with previous doses, the bivalent vaccine was generally better tolerated than previously received COVID-19 vaccines, with some minor differences between individuals with and without prior infection (figure 1C, p=0.032).

**Figure 1:**
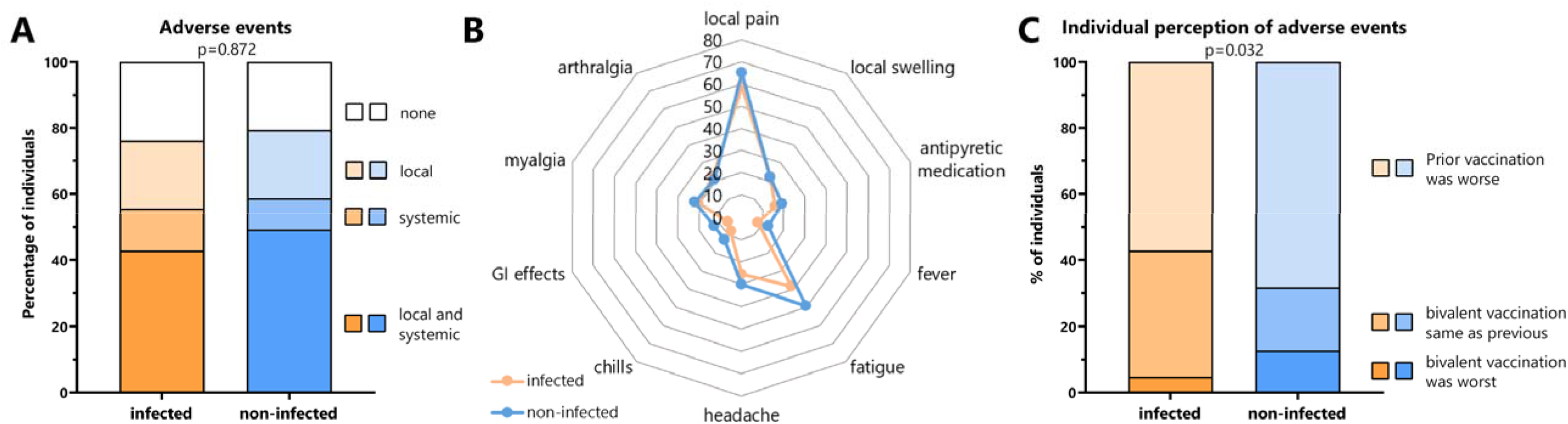
Adverse events after the bivalent vaccination. (A) Local and systemic adverse events in the first seven days after the bivalent vaccination were self-reported using a questionnaire. Shown is the percentage of individuals with (n=64) and without infection (n=63) who reported local or systemic adverse events or both. **(B)** The percentage of individuals with individual local or systemic adverse events and use of antipyretic medication. **(C)** Individual perception of relative severity of adverse events is shown based on whether individuals had felt more affected by the bivalent dose or by a vaccine dose administered before. Statistical analysis was performed by the Χ^2^ test.

### Lower levels of spike-specific IgG and neutralizing activity in non-infected individuals

A schematic overview of blood sampling before and after vaccination is shown in figure 2A. Both individuals with and without prior infection had detectable spike-specific IgG before vaccination at baseline with significantly higher median levels in previously infected individuals with hybrid immunity (1714 (IQR 1921) BAU/ml) than in non-infected (465 (IQR 816) BAU/ml, p<0.0001, figure 2B). The bivalent vaccine led to a significant increase in IgG-levels in both groups (p<0.0001). Although individuals with hybrid immunity reached significantly higher IgG-levels (7544 (IQR 5566) BAU/ml) than non-infected (5045 (IQR 4751) BAU/ml), the relative increase was significantly higher in individuals without prior infection (9.5-fold versus 4.8-fold, p<0.0001, figure 2C). As shown by a micro-neutralization assay, baseline neutralizing activity towards the authentic parental SARS-CoV-2 strain (D614G, FFM7) were significantly higher in previously infected than in non-infected individuals and significantly increased after vaccination by at least 3 log-levels in both groups (p<0.0001, figure 2D). Likewise, median titers after vaccination were also significantly higher in individuals with hybrid immunity (p<0.0001), and more than 50% of infected individuals reached neutralization titers above the upper limit of quantification. Compared to FFM7, baseline neutralizing activity towards the authentic Omicron subvariant BA.5 targeted by the vaccine, as well as BA.1 and BA.2 was generally lower with marked differences between previously infected and non-infected individuals (p<0.0001, figure 2D). In line with the fact that the majority of infections had occurred in the BA.2 era (table 1), individuals with hybrid immunity had higher median baseline neutralizing titers towards BA.2 than towards BA.1 or BA.5. Their vaccine-induced increase in neutralizing titers was more pronounced for BA.5 (by 3 log-levels) than for BA.1 or BA.2 (by ≥2 log-levels). Among non-infected individuals, the majority had negative baseline titers towards all Omicron subvariants, which increased by ≥4 log-levels not only for BA.5, but also for BA.1 and BA.2 (figure 2D). As shown by the correlation matrix in figure 2E, IgG levels and neutralizing antibody activity towards the various Omicron subvariants showed significant correlations in both individuals with and without previous infection.

**Figure 2:**
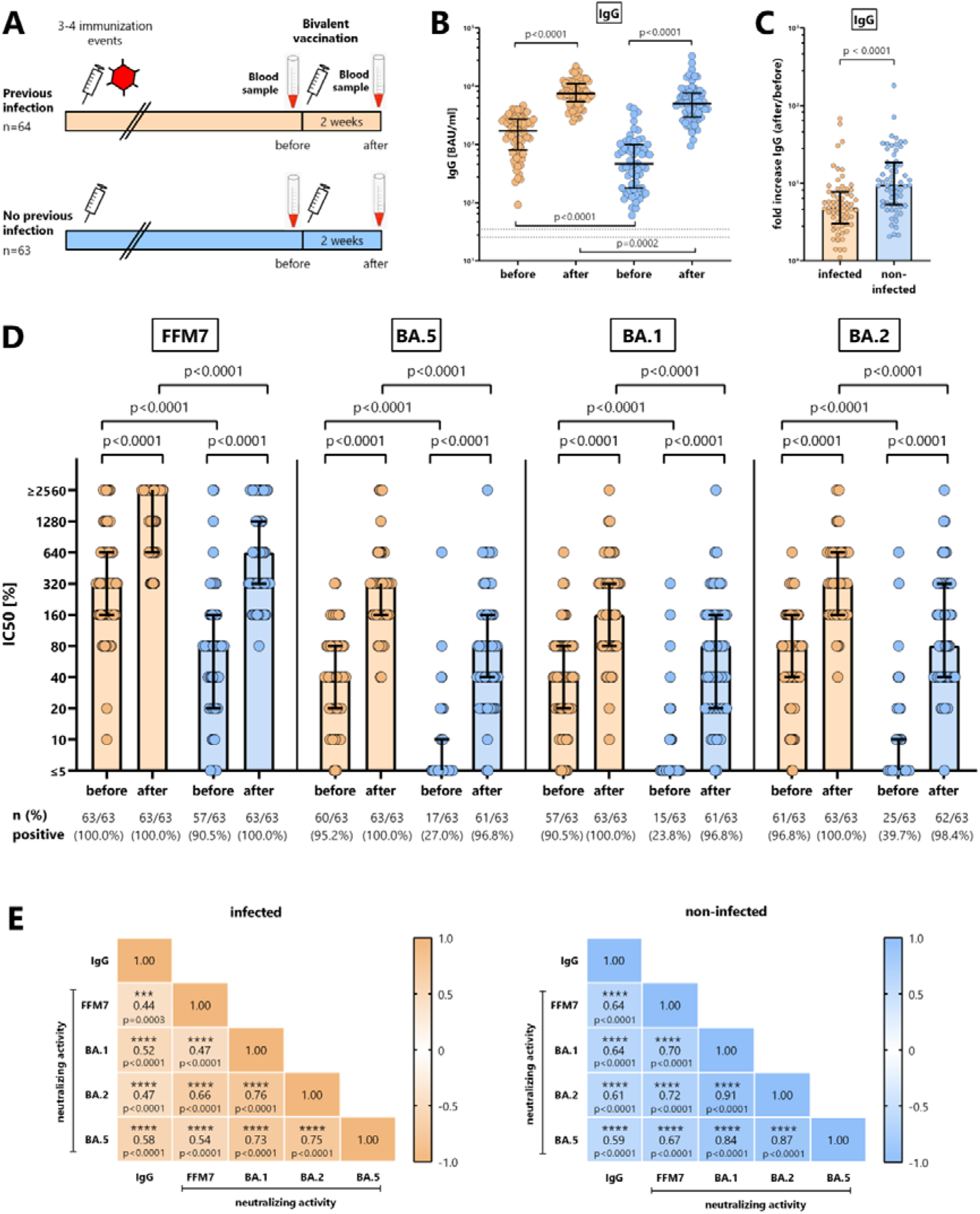
Spike-specific IgG and neutralizing antibodies before and after bivalent vaccination. (A) Schematic representation of study design and blood sampling. **(B)** Spike-specific IgG levels (in BAU/ml) towards the parental SARS-CoV-2 spike protein were determined from individuals with (orange symbols, n=64) and without prior infection (blue symbols, n=63) before and after bivalent vaccination. Statistical analysis was performed using the paired t-test (before/after) or the non-parametric Mann-Whitney test for between-group comparisons at baseline and after vaccination. **(C)** The fold increase in spike-specific IgG levels was determined for individuals with and without prior infection and compared using Mann-Whitney test. **(D)** Neutralizing activity of antibodies towards authentic parental SARS-CoV-2 (FFM7) and Omicron subvariants were determined in infected (n=63) and non-infected individuals (n=63) using a microneutralization assay, and differences were calculated using the paired t-test (before/after) or the non-parametric Mann-Whitney test for between-group comparisons at baseline and after vaccination. **(E)** Correlation matrix between IgG levels and neutralizing activities towards Omicron subvariants. Correlation coefficients were calculated according to two-tailed Spearman and displayed using a color code, and p-values (including stars denoting levels of statistical significance) are indicated. Bars indicate medians and interquartile ranges. SEB, *Staphylococcus aureus* Enterotoxin B.

### Quantitative similarities in spike-specific CD4 and CD8 T-cells towards parental spike and Omicron subvariants irrespective of infection history

Specific CD4 and CD8 T-cells towards the parental spike were quantified before and after vaccination. Specific T-cells were identified after stimulation with overlapping peptides followed by intracellular staining of IFNγ in CD69 positive CD4 and CD8 T-cells with a representative example shown in Extended Data figure 2. As shown in figure 3A, the vaccine induced a significant increase in both specific CD4 and CD8 T-cells (p<0.0001). Interestingly, unlike spike-specific antibodies, there was no difference between individuals with and without prior infection, which held true for both baseline levels and vaccine-induced levels of specific CD4 and CD8 T-cells (figure 3A). In contrast, changes in the magnitude of polyclonally stimulated CD4 and CD8 T-cells were less pronounced (figure 3B).

**Figure 3:**
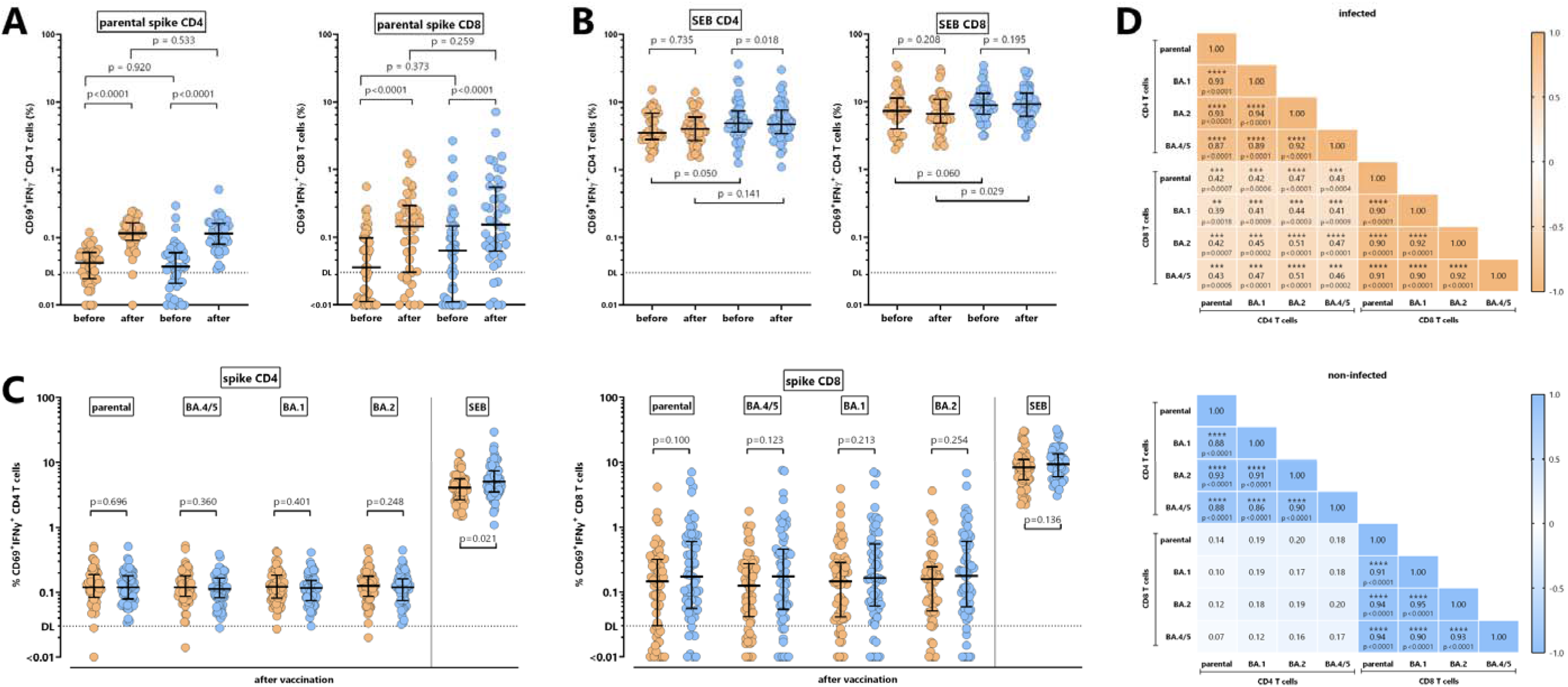
Spike-specific CD4 and CD8 T-cells towards parental spike and Omicron subvariants. (A) Specific CD4 and CD8 T-cells towards SARS-CoV-2 parental spike and (**B)** SEB-reactive CD4 and CD8 T-cells were quantified in subgroups of infected (orange symbols, n=44) and non-infected individuals (blue symbols, n=45) before and after bivalent vaccination. Reactive CD4 and CD8 T-cells were quantified after stimulation based on expression of CD69 and IFNγ. Statistical analysis was performed using the Wilcoxon matched pairs t-test (before/after) or the Mann-Whitney test for between-group comparisons at baseline and after vaccination. **(C)** CD4 and CD8 T-cells towards parental spike and towards the spike-protein of the Omicron subvariants BA.4/5, BA.1 and BA.2 were determined after bivalent vaccination (infected n=64 (n=63 for BA.2), non-infected n=63). SEB-reactive CD4 and CD8 T-cell levels were quantified as positive controls. Statistical analysis was performed using the Mann-Whitney test. Bars indicate medians and interquartile ranges. Stippled lines denote detection limits as defined in previous studies^18, 42^. **(D)** Correlation matrix of specific CD4 and CD8 T-cells towards the parental spike and spike of the Omicron subvariants in infected and non-infected individuals. Correlation coefficients were calculated according to two-tailed Spearman and displayed using a color code, and p-values (including stars denoting levels of statistical significance) are indicated. SEB, *Staphylococcus aureus* Enterotoxin B.

We also analyzed vaccine-induced CD4 and CD8 T-cells against the Omicron BA.4/5 spike targeted by the vaccine, and against the two other Omicron subvariants BA.1 and BA.2. As shown in figure 3C, vaccine-induced CD4 and CD8 T-cell levels against the Omicron variants were of similar magnitude as those against the parental spike. Moreover, unlike antibodies, specific CD4 and CD8 T-cell levels did not differ between individuals with and without prior infection (figure 3C). Within either CD4 or CD8 T-cells, there was a strong correlation in the percentages of T-cells towards parental spike and BA.1, BA.2 and BA.4/5. Interestingly, however, spike-specific CD4 and CD8 T-cells only correlated in individuals with previous infection, whereas no such correlation was found in non-infected individuals (figure 3D).

### Correlations between spike-specific IgG, neutralizing activity and cellular immunity

Individuals with and without prior infection differed in their correlation patterns of CD4 and CD8 T-cells with humoral immune response parameters. In individuals with prior infection, specific CD8 T-cell levels towards spike of the parental strain or of the Omicron subvariants BA.1, BA.2 or BA.4/5 correlated with IgG titers, and neutralizing activity towards the parental SARS-CoV-2, and in part towards Omicron BA.1 or BA.2, whereas specific CD4 T-cells did not show any correlation with humoral immunity (Extended Data Fig. 3A). In contrast, among individuals without prior infection, specific CD4 T-cell levels towards spike of the parental SARS-CoV-2 or of the Omicron subvariants BA.1, BA.2 or BA.4/5 correlated with IgG titers, and neutralizing activity towards the parental SARS-CoV-2, and BA.5, and in part towards BA.1 or BA.2 variants, whereas specific CD8 T-cells did not show any correlation with humoral immunity (Extended Data Fig. 3B).

### Phenotypical and functional similarities in spike-specific CD4 and CD8 T-cells towards parental spike and Omicron subvariants irrespective of infection history

Apart from quantitative analyses, spike-specific CD4 and CD8 T-cells were further characterized phenotypically for their expression of CTLA-4 as a marker for recent antigen encounter. As shown in figure 4A, CLTA-4 expression on spike-specific T-cells was significantly higher as compared to *Staphylococcus aureus* Enterotoxin B (SEB)-reactive CD4 and CD8 T-cells, which indicates that the effect is vaccine-specific. Moreover, CLTA-4 expression levels of vaccine-induced CD4 and CD8 T-cells reactive towards spike of the parental strain and the Omicron subvariants were similarly high irrespective of prior infection. Functional analysis of spike-specific CD4 and CD8 T-cell subpopulations with the ability to produce IFNγ, TNFα and IL-2 alone or in combination showed that the cytokine expression profile was similar for cells reactive towards parental strain and the three VOCs (figure 4B). Specific CD4 T-cells were predominantly multifunctional with the ability to simultaneously produce all tested cytokines, whereas specific CD8 T-cells were predominantly producing IFNγ and TNFα and largely lacked the ability to express IL-2. As with CTLA-4 expression, there was no difference in the cytokine profiles of CD4 and CD8 T-cells between individuals with and without prior infection (figure 4B).

**Figure 4:**
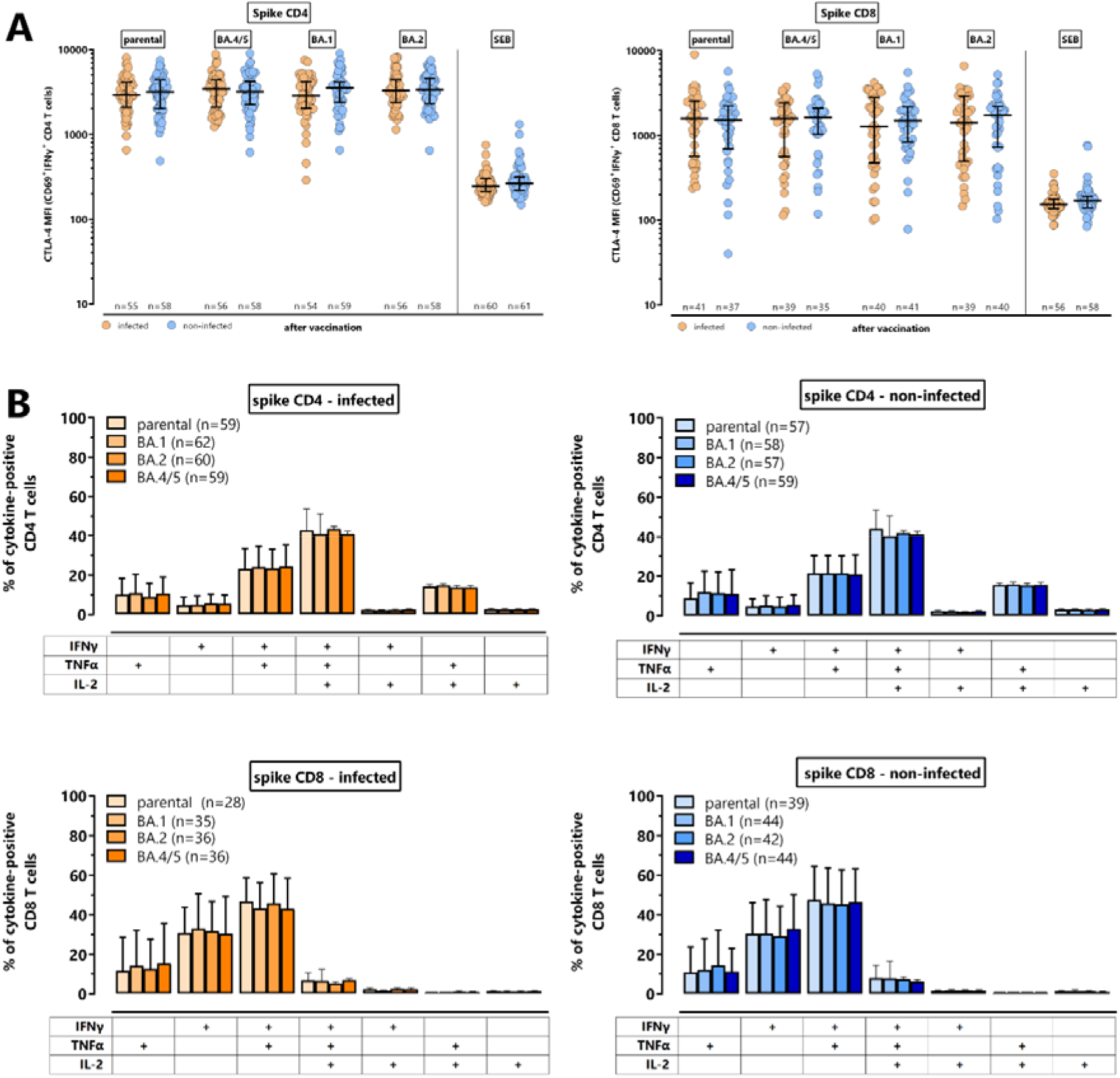
CTLA-4 expression and cytokine profile of spike-specific and SEB-reactive CD4 and CD8 T-cells. (A) Specific CD4 and CD8 T-cells towards the parental spike and Omicron subvariants BA.4/5, BA.1 and BA.2 as well as SEB-reactive CD4 and CD8 T-cells were analyzed for expression of CTLA-4, which is expressed as median fluorescence intensity (MFI). All samples were analyzed, but to ensure robust statistics, this analysis was restricted to samples with at least 20 CD69+IFNγ+ CD4 or CD8 T-cells (with sample size indicated in the figures). Bars represent medians with interquartile ranges. Differences between the groups were calculated using the Mann Whitney test. **(B)** Specific CD4 and CD8 T-cells towards the parental spike and Omicron subvariants BA.4/5, BA.1 and BA.2 were analyzed for expression of IFNγ, TNFα and IL-2 alone or in combination after Boolean gating. This allowed distinction of seven subpopulations expressing three, two or a single cytokine. All samples were analyzed, but to ensure robust statistics, only samples with at least 30 cytokine-expressing spike-specific CD4 or CD8 T-cells after normalization to the negative control stimulation were considered (with sample size indicated in the figures). Bars represent means and standard deviations, and differences between the groups were analyzed using the Kruskal-Wallis with Dunn‘s post test. CTLA-4, cytotoxic T-lymphocyte antigen 4; SEB, *Staphylococcus aureus* Enterotoxin B.

### Lower vaccine-induced neutralizing activity, and spike-specific CD4 T-cell levels in non-infected individuals with subsequent breakthrough infection

All individuals were followed up for development of a breakthrough infection after a median observation time of 146 (IQR 10) days using a questionnaire (figure 5A). 25/126 (19.8%) individuals developed a breakthrough infection at a median of 129 (IQR 65) days after the bivalent vaccination, which occurred more often in individuals without prior infection (16/62 (25.8%)) than in individuals with a previous infection (9/64 (14.1%), p=0.045, figure 5B and Extended Data Fig. 4). The incidence rates did not differ significantly (106 (95% CI 49-202) cases/100.000 person-days and 194 (95% CI 111-315) cases/100.000 person-days in individuals with and without prior infection, p=0.145). Most individuals only had mild or moderate symptoms, none of which required hospitalization. Although the duration of these symptoms did not differ between individuals with and without prior infection, symptoms lasting 7 days or more were numerically more frequent among individuals without previous infection (Extended Data Fig. 4).

**Figure 5:**
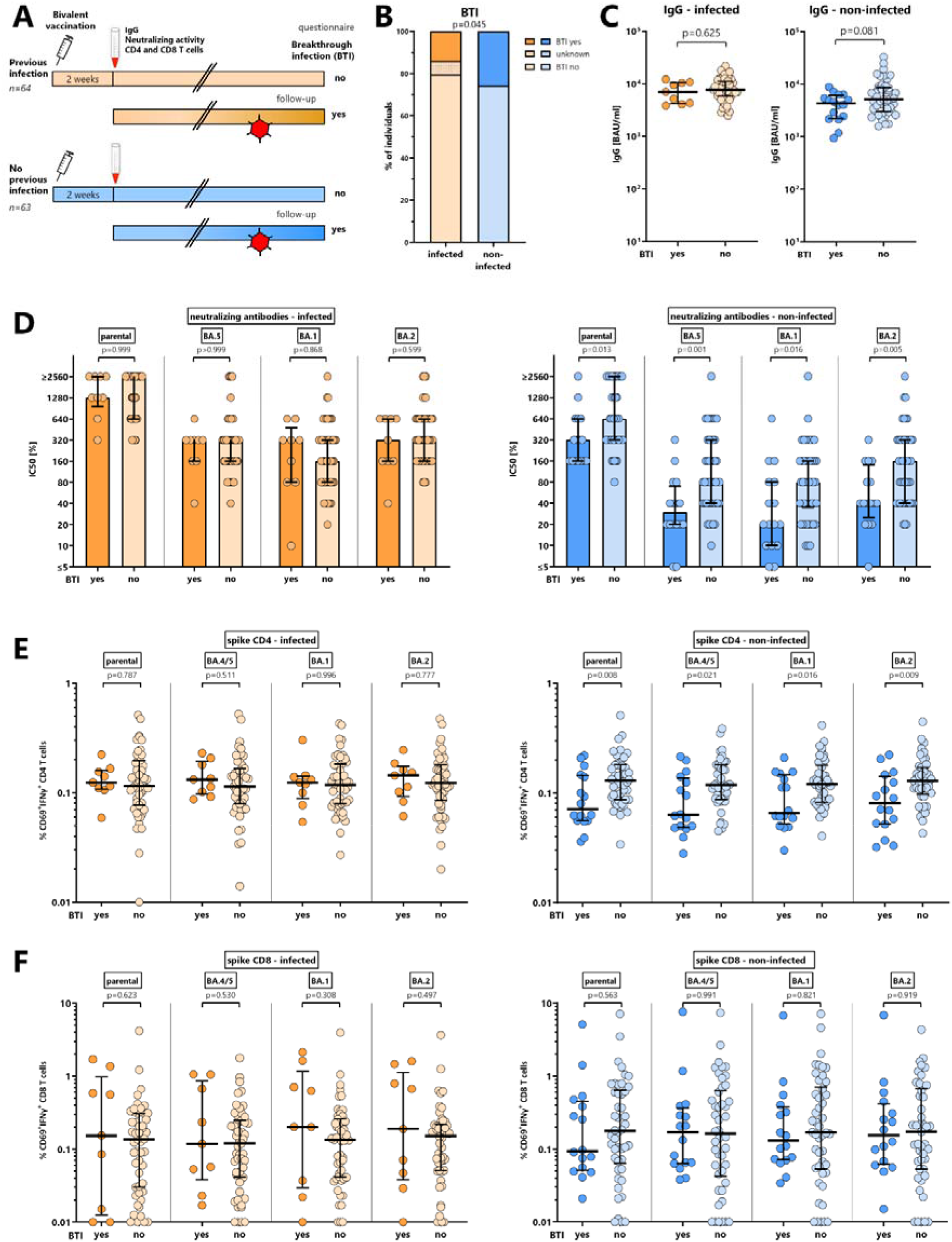
Vaccine-induced humoral and cellular immunity in individuals with and without breakthrough infections. (A) Schematic outline of the study design. All study participants were followed up until March 2023 for development of breakthrough infections based on self-reporting using a questionnaire. Immune parameters in the following panels refer to results two weeks after the bivalent vaccination. **(B)** Percentage of breakthrough infections (BTI) in individuals with (orange, n=64) and without (blue, n=62) prior infection. Bivalent vaccine-induced spike-specific **(C)** IgG levels, **(D)** neutralizing antibody activity, **(E)** CD4 T-cells, and **(F)** CD8 T-cells in individuals with and without prior infection, stratified for individuals with and without subsequent breakthrough infection. Statistical analysis was performed using the Mann-Whitney test. BTI, breakthrough infection.

We then analyzed whether the levels of humoral and cellular immunity determined two weeks after vaccination differed in individuals with and without subsequent breakthrough infection, which was not the case for IgG levels (figure 5C). Interestingly, however, both neutralizing activity and spike-specific CD4 T-cell levels towards the parental strain and all Omicron VOCs were significantly lower in previously non-infected individuals who developed a breakthrough infection, whereas no such difference was found for spike-specific CD8 T-cells (figure 5D-F, right panels). In contrast, vaccine-induced neutralizing antibody activity and spike-specific CD4 or CD8 T-cells did not differ among previously infected individuals with or without subsequent infection (figure 5D-F, left panels).

## Discussion

In this observational study we show that the bivalent Comirnaty Original/Omicron BA.4/5 vaccine strongly induced specific IgG levels, neutralizing activity, and specific CD4 and CD8 T-cells, which were not only directed towards the spike proteins of the parental SARS-CoV-2 and BA.4/5 strains targeted by the vaccine, but also towards Omicron subvariants BA.1 and BA.2. Approximately 50% of our study participants had hybrid immunity based on prior infection, and the vaccine was well tolerated in both groups. However, individuals with and without hybrid immunity showed marked differences in the induction of both IgG levels and neutralizing antibody titers, whereas vaccine-induced T-cell levels were induced to a similar extent. Finally, we show that individuals without prior infection more frequently developed a SARS-CoV-2 breakthrough infection after vaccination. As most striking finding, previously non-infected individuals who subsequently developed breakthrough infections not only had significantly lower neutralizing activity, but also lower spike-specific CD4 T-cell levels after vaccination. This demonstrates that the magnitude of vaccine-induced neutralizing activity and CD4 T-cell response may serve as a correlate for protection among individuals without prior infection.

Vaccine-induced IgG levels and neutralizing antibody activity towards the various Omicron subvariants showed significant correlations in both individuals with and without previous infection. Higher antibody levels and neutralizing activity towards the ancestral spike as compared to the Omicron subvariants indicate some extent of immune imprinting and are in line with recent reports on neutralizing antibodies after vaccination with a bivalent BA.1^6^ or BA.4/5 vaccine^7–10^. However, these studies were of small sample size^7–10^, reported aggregated data for vaccinated individuals with and without prior infection^9, 10^ or did not report pre-vaccination data^8, 10^ to appreciate dynamic changes in neutralizing activity before and after vaccination in individuals with and without prior infection. In general, immune imprinting by previous exposures with the monovalent vaccine may account for the relative dominance of neutralizing activity towards the parental spike as compared to the Omicron subvariants^15, 16^. Among individuals with prior infection, it was interesting to note that baseline immunity against the ancestral SARS-CoV-2 and the BA.2 variant was higher than against BA.1 or BA.5. This well reflects previous exposure to these antigens by vaccination and subsequent BA.2 infection, which was the most prevalent infection strain in our study participants. Despite some extent of imprinting, the most pronounced increase was found for antibody titers towards the two SARS-CoV-2 spike variants against which the vaccine was also directed. Moreover, consistent with infection representing one more immunization event, individuals with prior infection had both higher baseline titers and reached higher absolute levels of vaccine-induced humoral immunity. As with other vaccines such as influenza^17^, higher levels of baseline antibody titers were associated with a less pronounced relative increase after vaccination. In this regard, the relative increase in vaccine-induced humoral immunity was more pronounced in individuals without prior infection, which was particularly striking for neutralizing titers towards the Omicron subvariants, which were negative in most cases at baseline and increased by 4 log-steps after vaccination. It therefore seems that the additional bivalent vaccine dose may confer a stronger benefit for previously non-infected individuals.

Unlike antibody levels, baseline frequencies of spike-specific CD4 or CD8 T-cells were equally low in individuals with and without prior infection and both groups showed a similar increase after bivalent vaccination. In general, vaccine-induced T-cells were largely polyfunctional with high CTLA-4 expression levels as sign of recent antigen encounter, and CD8 T-cell levels were higher than CD4 T-cells. Interestingly, spike-specific CD4 and CD8 T-cell levels only correlated in individuals with previous infections, whereas no such correlation was observed among non-infected individuals. This may suggest that the potent induction of both CD4 and CD8 T-cells by natural infection^18^ may allow for a more uniform expansion of both T-cell populations after re-challenge with the vaccine. Among each population of CD4 or CD8 T-cells, a strong correlation and striking similarity of vaccine-induced T-cells levels towards the parental spike and spike of all tested Omicron subvariants was found. This is in line with results after monovalent vaccination^19, 20^, and indicates substantial cross-reactivity of T-cells with little evidence of immune escape. In general, evolution of immune escape mutants affecting T-cells is less frequent due to the diversity of MHC alleles in the human population. Cross-reactivity on the T-cell level may on one hand result from the fact that more than 86% of class I and 72% of class II epitopes of the Omicron spike are fully conserved, and epitope mutations do not necessarily preclude T-cell recognition^19^. On the other hand, reactive T-cells have been shown to be directed towards non-mutated regions of the spike protein^21^. Data on the induction of T-cells after bivalent BA.4/5 vaccination are scarce. The marked increase in vaccine-induced T-cell levels in our study cohort contrasts with recent findings among 18 individuals where the bivalent vaccine did not lead to a substantial augmentation in cellular immunity^9^. The reason for this difference is unclear but it seems that T-cell levels at baseline were higher than in our study, which may be due to a smaller interval between the previous immunization event and the bivalent vaccination. As the authors speculate that the majority of their participants had hybrid immunity due to a recent BA.5 infection during the summer and fall of 2022^9^, this may have boosted specific T-cell immunity already prior to vaccination thereby diminishing a further booster effect by the vaccine.

Data on the effectiveness of bivalent vaccines in individuals with and without prior infection are limited, but previous studies have shown that hybrid immunity conferred stronger protection from Omicron infection than monovalent vaccine-induced immunity alone^22–25^. Early estimates of bivalent vaccines in general suggest a high effectiveness against hospitalization and death when given as additional dose or as alternative to the monovalent dose^26–28^. As with a recent study after bivalent BA.1/2 vaccination^6^, our study after bivalent BA.4/5 vaccination also showed a lower percentage of breakthrough infections among individuals with a previous infection, which mostly occurred during the BA.2 wave in Germany. So far, data linking results on bivalent vaccine-induced humoral and cellular immunity to subsequent breakthrough infections are lacking. We now show that vaccine-induced humoral and cellular immunity among previously infected individuals had no value in predicting additional infections. This may be due to the fact that previously infected individuals had higher levels of IgG and neutralizing activity as compared to previously non-infected, and may indicate that other components of the adaptive immune response such as local immunity in the respiratory tract may play an additional role towards protection^29^. Interestingly, among previously uninfected individuals, those with subsequent breakthrough infections had significantly lower levels of vaccine-induced neutralizing activity and CD4 T-cells, which suggests that these parameters may serve as a correlate of protection in individuals without hybrid immunity. This is conceivable, as helper CD4 T-cells are instrumental in supporting induction of antibodies, and both parameters showed a significant correlation in non-infected individuals only. Low neutralizing antibody levels were already identified before to be predictive of both infection and severity of disease^24, 30, 31^, which was also shown in a prospective household study among contacts without previous infections during the Delta wave that also analyzed the role of T-cells^32^. Unlike in our study, T-cell activity determined by an ELISPOT assay did not have any predictive value, but given that CD4 and CD8 T-cells were not distinguished, this may have masked a protective effect of individual T-cell subpopulations. Future studies are needed to further corroborate the particular role of CD4 T-cells as correlate of protection, which may be of particular interest for individual risk assessment and guidance of booster vaccination for vulnerable patient populations with limited antibody responses^12, 14, 33^.

Our study has some limitations. Analyses of breakthrough infection relied on self-reporting which may have underestimated the true infection rate and we were unable to address correlations with severe disease as none of the breakthrough infections required hospitalization. Moreover, we have not performed follow-up analyses to assess peri-infection immune responses. Nevertheless, together with the known waning of humoral immunity following vaccination^34–36^, lower post-vaccine responses may have more rapidly decreased beyond a level of protection on follow-up. Due to the real-world setting, we were unable to study effects of a monovalent booster dose, as the bivalent vaccination was preferentially recommended at the start of our study. A strength of our study is the considerably large sample size with similar numbers of individuals with and without history of prior infection, which allows evaluating the effect of prior infection on immunogenicity and on protection from further infection. All vaccinations were carried out in the same time frame in the same region with similar circulating variants before and after vaccination. Moreover, we included analyses of both antibodies and cellular immune responses. Finally, the considerably high infection rate in the Omicron BA.5 wave of around 20% allowed a better evaluation of individual participant-based immunological correlates of protection even from small sample sizes.

In conclusion, we have shown that the bivalent BA.4/5 vaccine was well tolerated and was strongly immunogenic with marked difference in individuals with and without prior infection. This is the first study after bivalent vaccination that has shown a link between the magnitude of vaccine-induced neutralizing antibodies and CD4 T-cells and protection from infection among individuals without prior infection. Together with the individual history of infection and vaccination, the magnitude of neutralizing antibody titers and CD4 T-cells will allow refinement of the individual risk for infection, and may therefore be of relevance for future vaccine strategies.

## Online methods

### Study participants and study design

In an observational study, immunocompetent individuals receiving a bivalent COVID-19 vaccine (Comirnaty Original/Omicron BA.4-5, BioNTech/Pfizer) were enrolled in the study between 28th of September and 14th of December 2022 as per German recommendations. Participants were recruited either from the Saarland University Medical Center (Homburg, Germany) or from a public vaccination campaign (St. Ingbert, Germany). A heparinized blood sample was scheduled before and 13-18 days after vaccination to determine differential blood counts and immunogenicity. In addition, all participants reported their history of SARS-CoV-2 infection and COVID-19 vaccination, and vaccine-related adverse events in the first week after vaccination, using a standardized questionnaire. Finally, all individuals were interrogated in March 2023 for potential development of a breakthrough infection after the bivalent vaccination (see Extended Data Fig. 1). The study was approved by the ethics committee of the “Ärztekammer des Saarlandes” (reference 76/20 including amendment), and all individuals gave written informed consent.

### Preparation of UV-inactivated viral strains

Sample inactivation for further processing and normalization was performed as described previously^20, 37^. Description of SARS-CoV-2 isolates are given in the supplementary methods section. The following SARS-CoV-2 isolates were used in this study: Parental (SARS-CoV-2 B.1 FFM7/2020, GenBank ID MT358643), BA.1 (SARS-CoV-2 B.1.1.529 FFM-SIM0550/2021 (EPI_ISL_6959871), GenBank ID OL800702), BA.2 (SARS-CoV-2 BA.2 FFM-BA.2-3833/2022, GenBank ID OM617939), BA.5 (SARS-CoV-2 BA.5 FFM-BA.5-501/2022, GenBank ID OP062267)^3, 38–41^.

### Quantitation of SARS-CoV-2 specific CD4 and CD8 T-cells

SARS-CoV-2 spike-specific T-cells were measured after a 6h stimulation of heparinized whole blood with overlapping peptide pools derived from the S1 and S2 domain of the parental SARS-CoV-2 spike protein exactly as previously described^42, 43^. In addition, peptide pools from the BA.1, BA.2 and BA.4/5 spike protein were used (jpt, Berlin, Germany). Spike-specific CD4 or CD8 T-cells were identified by co-expression of CD69 and IFNγ and further characterized for expression of CTLA-4, and of the cytokines IL-2 and TNFα. Specific CD4 or CD8 T-cell levels ≥0.03% after subtraction of control stimulations were scored as positive as previously established from cohorts of infected and vaccinated individuals^18, 42^. Analyses were carried out on a FACS Canto II using Diva software (BD, Heidelberg, Germany). The gating strategy is shown in Extended Data Fig. 5, antibodies used in the study are listed in Extended Data Fig. 6.

### Quantitation of SARS-CoV-2 specific IgG and neutralizing activity

The amount of SARS-CoV-2 spike-specific IgG antibodies was determined using an ELISA (SARS-CoV-2-QuantiVac, Euroimmun, Lübeck, Germany) as described before^42, 43^. Antibody binding units (BAU/ml) <25.6 were scored negative, ≥25.6 and <35.2 were scored intermediate, and ≥35.2 were scored positive. SARS-CoV-2 specific IgG towards the nucleocapsid protein (NCP) were quantified using the anti-SARS-CoV-2-NCP-ELISA (Euroimmun). The *in vitro* neutralizing activity of the antibodies was measured using a micro neutralization assay with A549-AT-cells^44^ and authentic parental SARS-CoV-2 (FFM7, D614G) and the Omicron subvariants BA.1, BA.2, and BA.5 as described before^3, 20^.

### Statistical analysis

The Mann-Whitney test or the paired t-test was used to analyze differences between non-parametric data such as blood cell populations, T-cell and antibody levels, and CTLA-4 expression. The Kruskal-Wallis test was performed for paired analyses of the cytokine-profiles of CD4 and CD8 T-cells towards parental spike and spike of the Omicron subvariants. Age was analyzed using a non-paired t-test. Categorial analyses were performed using the Fisheŕs test or X^2^ tests. Correlations between the immunological parameters were analyzed using a correlation matrix according to Spearman. A p-value <0.05 was considered statistically significant. Analysis was carried out using GraphPad Prism 9.5.1 software (GraphPad, San Diego, CA, USA).

## Data Availability

All data produced in the present study are available upon reasonable request to the authors

## Acknowledgements

The authors thank all participants to this study, and the team of the occupational health care center at Saarland University Medical Center and the CDU Rentrisch for their support in enrolling study participants. Expert technical assistance by Christiane Pallas is acknowledged. Financial support was provided in part by the State chancellery of the Saarland to M.S. Moreover, the work was in part supported by the cluster project ENABLE, the Innovation Center TheraNova, and the LOEWE Priority Program CoroPan funded by the Hessian Ministry for Science and the Arts (HMWK) to M.W.

## Author Contributions

R.U., T.S. and M.S. designed the study; R.U., S.B., M.W., T.S. and M.S. designed the experiments, R.U., S.B., V.K., A.W., S.M., F.H., A.A.-O., M.W., and C.G. performed experiments; A.A.-O., S.S., C. B., S.L.B., B.C.G., and U.S. contributed to study design, patient recruitment, and clinical data acquisition. R.U., S.B., T.S. and M.S. performed statistical analysis. R.U., T.S., U.S., M.W., and M.S. supervised all parts of the study, and performed analyses; M.S. wrote the manuscript. All authors approved the final version of the manuscript.

---

**Extended data figure 1:**
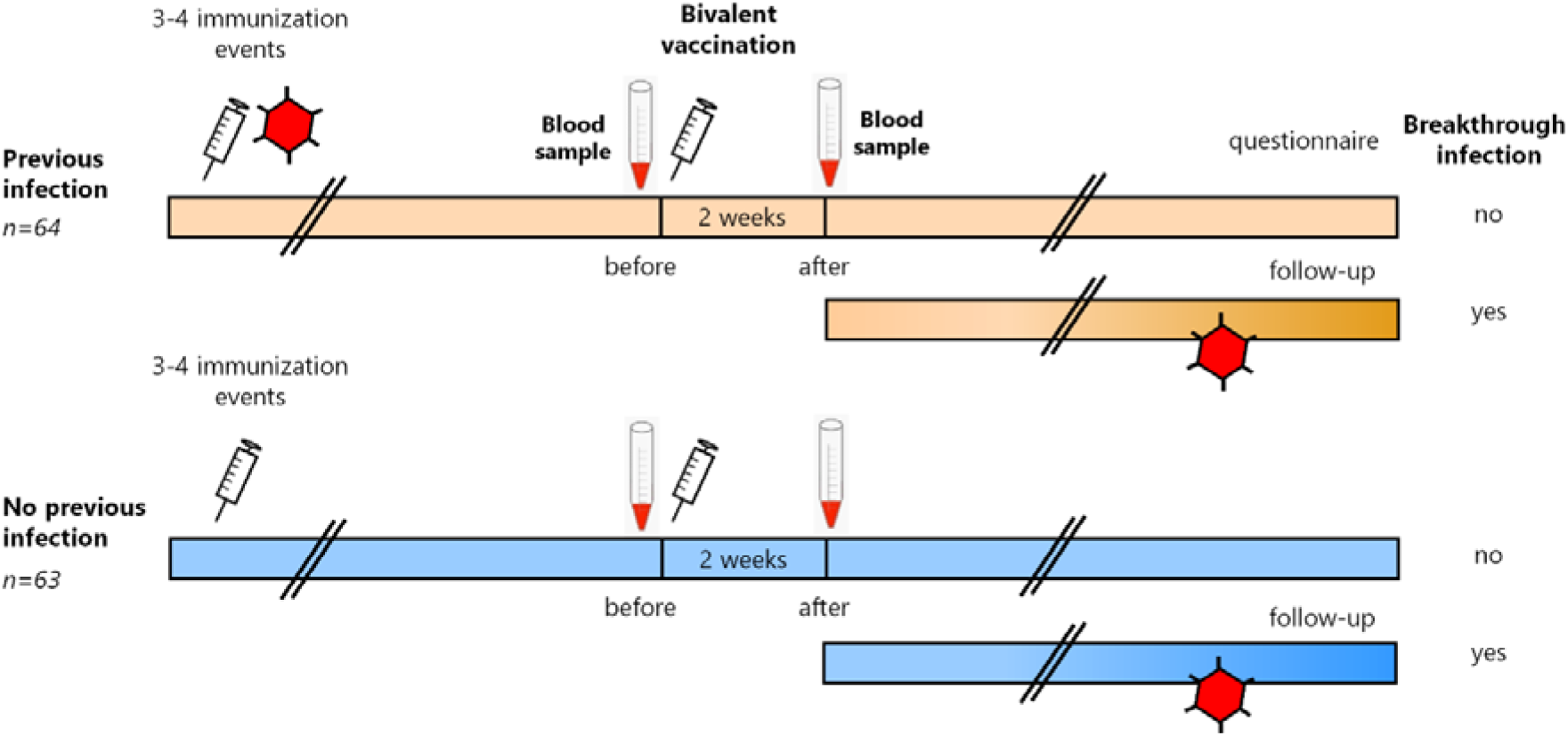
Schematic representation of study design. Individuals after 3-4 immunization events with (orange symbols, n=64) and without prior infection (blue symbols, n=63) were recruited. Blood sampling was performed before and after bivalent vaccination. All individuals were followed up for occurrence of breakthrough infection using a questionnaire.

**Extended data figure 2.**
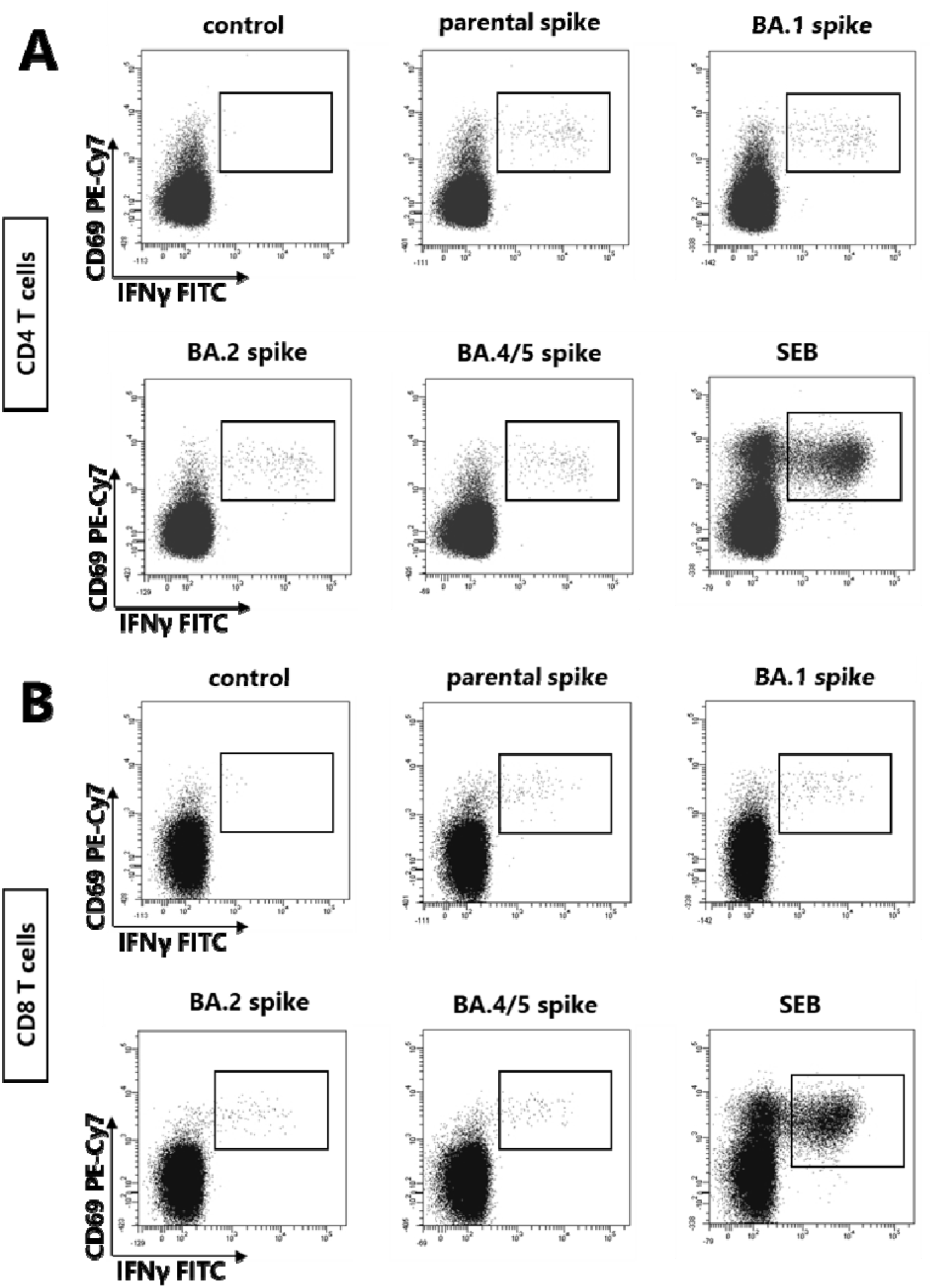
: Representative analysis of spike- and SEB-reactive CD4 and CD8 T cells. Stimulations of whole blood from a previously infected individual were carried out with negative control (DMSO diluent) as well as with overlapping peptide pools derived from spike of the parental parental SARS-CoV-2 and Omicron subvariants BA.1, BA.2, BA. 4/5 or with SEB as a polyclonal stimulus. **(A)** CD4 and **(B)** CD8 T cells co-expressing the activation marker CD69 and the cytokine IFNγ are shown. Boxes were used as gates to quantify the percentage of CD69^+^/IFNγ^+^ CD4 or CD8 T cells; SEB, *Staphylococcus aureus* Enterotoxin B.

**Extended data figure 3:**
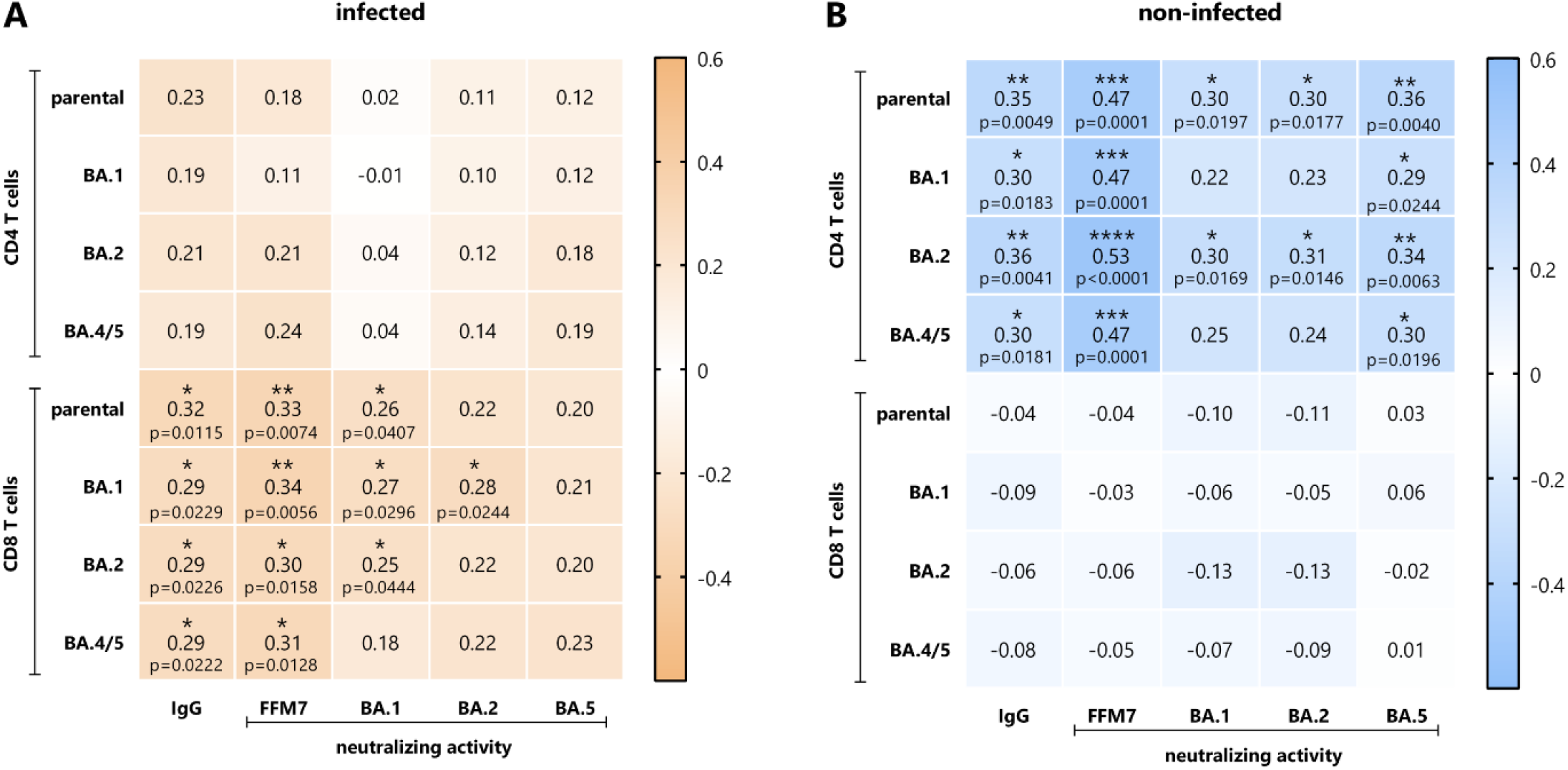
Correlation between vaccine-induced cellular and humoral immunity. Correlation matrix between vaccine-induced spike-specific CD4 or CD8 T-cells with IgG levels and neutralizing activities towards parental SARS-CoV2 and Omicron subvariants in individuals with **(A)** and **(B)** without prior infection. Correlation coefficients were calculated according to two-tailed Spearman and displayed using a color code, and p-values (including stars denoting levels of statistical significance) are indicated.

**Extended Data Fig. 4.**
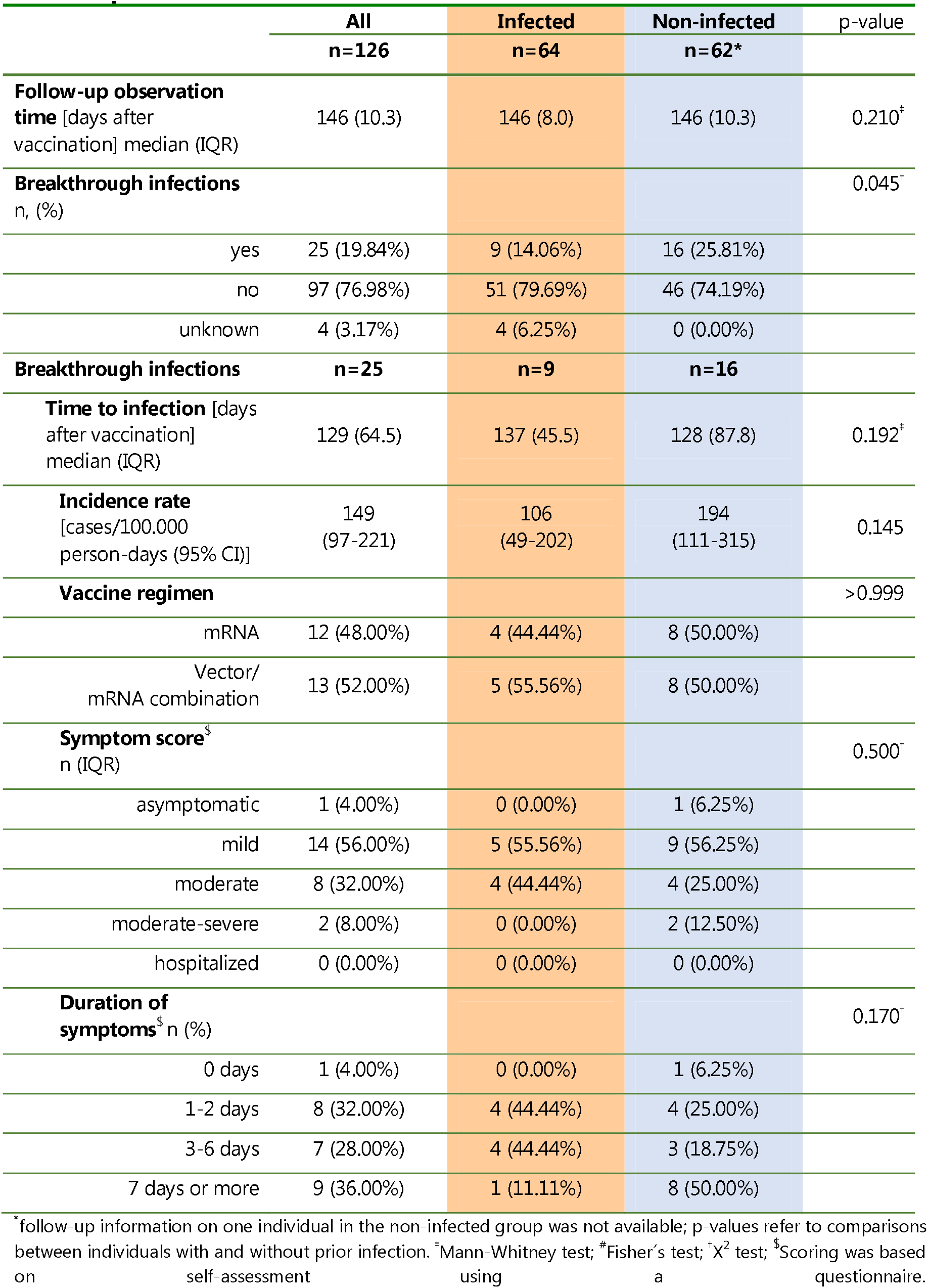
Clinical characteristics of breakthrough infections in individuals with and without infection prior to bivalent vaccination

**Extended data figure 5:**
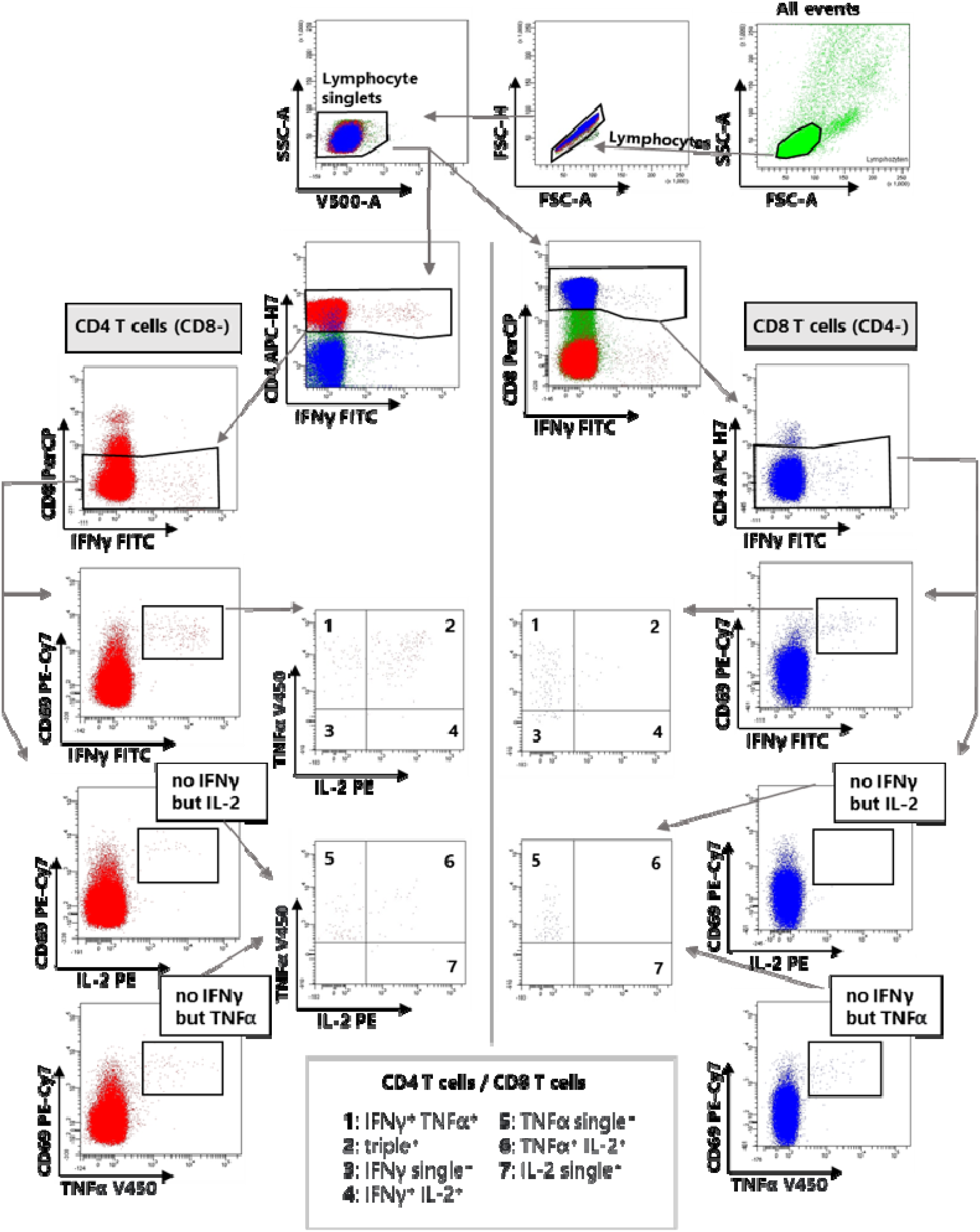
Gating strategy for identification of antigen-specific CD4 and CD8 T cells after stimulation. Lymphocytes were identified among total events by backgating of CD4 and/or CD8 positive cells combined with signals for size (FSC) and granularity (SSC). Hight and area signals of FSC were used to exclude doublets. The gating strategy to identify CD4 T cells (left side) or CD8 T cells (right side) co-expressing the activation marker CD69 and the cytokines IFNγ, IL-2 or TNFα are shown. Boxes were used as gates to quantify the percentage of CD69^+^/IFNγ^+^ CD4 or CD8 T cells. Moreover, Boolean gating for cytokine profiling is shown that was used to identify subpopulations of CD4 or CD8 T cells expressing all three cytokines (triple^+^), two cytokines or one cytokine only.

**Extended Data Fig. 6.**
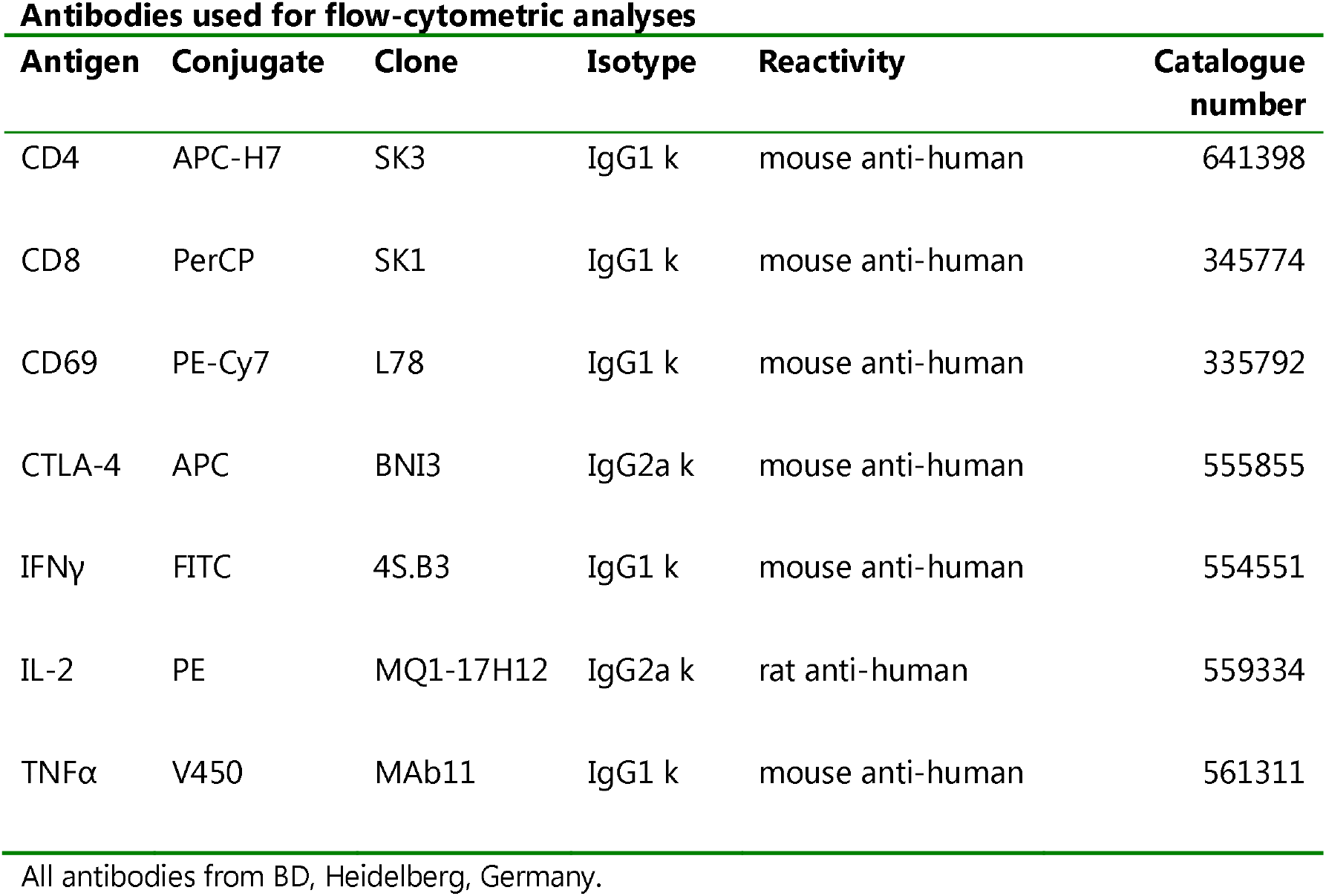
Antibodies used for flow-cytometric analyses

